# Examining the Implementation Process and Experience of health facility Autonomy Reforms in Kenya: A mixed methods study of counties in Kenya

**DOI:** 10.64898/2026.04.22.26351442

**Authors:** Anita Musiega, Jacinta Nzinga, Beatrice Amboko, Harrison Ochieng, Beryl Maritim, Rose Muthuri, Rahab Mbau, Benjamin Tsofa, Peter Mugo, James Bukosia, Elizabeth Wangia, Khatra Ali, Robert Rapando, Joy Mugambi, Stephen Wandei, Victor Tole, Beffy Vill, Michael Dramiga Obanda, Lidya Munteyian, Ethan Wong, Caitlin Mazzilli, Wangari Ng’ang’a, Anne Musuva, Felix Murira, Ileana Vilcu, Matt Boxshall, Nirmala Ravishankar, Edwine Barasa

## Abstract

**Background:** Kenya’s facility autonomy reforms are intended to improve health system equity, efficiency, and responsiveness to community needs by shifting decision-making to the frontline. This study evaluates the implementation process and experience of facility autonomy reforms in Kenya post devolution of health services.

**Methods:** We conducted a concurrent mixed methods study of counties (n=6) in Kenya, selected based on their implementation of facility financial autonomy reforms as of June 2023. For the quantitative aspect, we assessed 141 randomly selected public health facilities across all levels of service provision. We then did a descriptive analysis to measure the level and perceptions of autonomy. For the qualitative aspect, we reviewed documents and interviewed purposively selected stakeholders (n=71) involved with autonomy reforms at national, county, and facility levels, cutting across health, finance, legal, political and community actors. We analyzed the transcripts thematically using NVivo 12.

**Results:** The emergence of the FIF reforms in Kenya was driven by the convergence of political, technical, and public needs. While counties have developed their own facility autonomy laws to fit local contexts, some provisions are not fully aligned with the national legislation. Some aspects of both the county specific and national laws are not implemented. These include allocation of matching funds from the exchequer and reimbursing facilities for expenses incurred from providing care to indigents and for unpaid bills. The implementation of autonomy also varies, with some aspects partially or not implemented.

Autonomy reforms have contributed to improved decision-making, staff satisfaction, availability of essential medicines, and facility maintenance. However, challenges have emerged, including the failure of counties to provide matching funds, which disproportionately affects lower-level facilities that do not generate revenue. Additionally, the absence of waiver repayment mechanisms has led to inequities, and the risk of increased service costs threatens financial accessibility for marginalized populations.

**Conclusion:** Facility autonomy reforms support people-centered decision-making and aligns with PHC principles. While these reforms hold promise for improving service delivery and access, their success depends on complementary measures such as sustainable funding mechanisms and stronger protections for vulnerable populations.

## INTRODUCTION

Decentralization aims to improve health system performance by bringing services closer to communities, enhancing participation, accountability, responsiveness, equity, and efficiency (1,2). It involves shifting decision-making from a central level to local governments or health facilities (2). Health facility autonomy, a pathway to operationalize decentralization in health, involves granting facility management increased authority to make decisions over finances, procurement, human resources, operations, and strategy (3,4).

Literature highlights that limited autonomy compromises health facility performance. It disempowers facility managers from effectively setting priorities, reduces the responsiveness of facilities to population health needs, causes bureaucratic delays, demotivates staff, and weakens health facility management and accountability (3–9). Evidence on the impact of health facility autonomy on facility performance is mixed, with studies showing improved facility performance in some areas but no=t others following autonomy reforms (4,10,11). This mixed impact could be due to how the autonomy reforms are implemented. This study examines the implementation processes and experiences of health facility autonomy reforms within Kenya’s devolved health system to better understand their effects on service delivery and outcomes and inform the ongoing reforms in Kenya as well as other LMICs.

In 2013, Kenya adopted a devolved system of governance, creating 47 county governments responsible for service delivery, including health, while the national Ministry of Health retained policy oversight. (12). Coordination among the counties is facilitated by the Council of Governors (COG), a caucus comprising all 47 governors. Health service provision is organized into six levels and four tiers of care - community health services (Level 1), Primary healthcare services (level 2 and 3), Primary referral facilities (level 4 and 5) and tertiary referral facilities (level 6). County public health facilities are financed through county government budget allocations, social health insurance reimbursements, donor funds, and out of pocket payments. The flow and management of financing to government facilities has been a key factor influencing the extent to which facilities have autonomy over their decisions.

Devolution in Kenya was accompanied by the introduction of a new public finance management framework, with the enactment of the Public Financial Management (PFM) Act, 2012. While public health facilities had increased financial and operational autonomy prior to devolution, this was reduced post-devolution, for most of the counties (3,4,13,14). The PFM act 2012 required that all revenues that are collected or received at the county level, including by public health facilities are transferred to the county revenue fund (CRF) account. Although the PFM Act gave provisions for retention of funds in some entities, this was rarely taken to consideration. Public hospitals lost both financial and operational autonomy, leading to multiple challenges including late payment of salaries for casuals, and decreased availability of medicine (13,14). Lower-level facilities (level 2 and 3) retained access to revenue from donor funds such as the Danish International development Agency (DANIDA) fund and national conditional grants such as the user fee forgone fund. For level 2 and 3 facilities, some lost control over revenue generated from social health insurance reimbursements while some retained partial control for all or some revenue streams from social health insurance, likely because the revenue they generated was not significant.

Challenges linked to limited health facility autonomy prompted some Kenyan counties to introduce autonomy reforms. These efforts were marked by fragmented legislation, with counties creating their own legal frameworks. To guide this process, in 2022, the COG developed a model Facility Improvement Financing (FIF) law that advocated for facility autonomy. However, the model law saw uneven adoption. In 2023, the national Ministry of Health adapted the model law to form a national FIF Act as part of broader health system reforms—including primary care network reforms, social health insurance reforms, and digital health reforms. Although the national law was to supersede the county laws, anecdotal evidence suggests that it was disempowered as it doesn’t provide for regulations, therefore counties continue to use their county specific laws. This study explores the implementation experience of health facility autonomy in Kenyan counties within this evolving policy context.

## METHODS

### Conceptual Framework

We designed this study as a process evaluation (15), focusing on the emergence, fidelity, and implementation experience of the FIF reforms (Musiega et al., 2024) .

### Study design

We conducted a concurrent mixed methods study of counties in Kenya. The qualitative interviews were done between February 2024 and March 2025 while the quantitative assessments were done between October and December 2023.

### Study Setting

We collected data at the national level, and in six out of the 47 counties in Kenya. We selected counties with autonomy reforms for more than 1 year as of June (2023) Kajiado (2021), Kisumu (2022), and Nakuru (2013) and those newly implementing the reforms, starting July 2023 (Migori, Kakamega) and October 2023 (Lamu) to allow for exploration at different implementation stages where the effects and challenges may be different (16) (Table 1 ) .

**Table 1:** Characteristics of study counties.

| County | Population (2019) | %population in the lowest wealth quintile (17) | Total Public Facilities in county(18) |
| --- | --- | --- | --- |
| Kisumu | 1,155,574 | 15.0 | 146 |
| Nakuru | 2,162,202 | 12.3 | 184 |
| Kajiado | 1,117,840 | 19.9 | 107 |
| Kakamega | 1,867,579 | 17.5 | 197 |
| Migori | 1,116,436 | 31.8 | 184 |
| Lamu | 143,920 | 49 | 35 |
| National estimate | 47,564,296 | 20 | 14,795 |

### Study Participants

#### Qualitative Study Participants

We purposively selected and interviewed participants involved in the formulation and implementation of health facility autonomy reforms across different levels (Table 2).

**Table 2.**
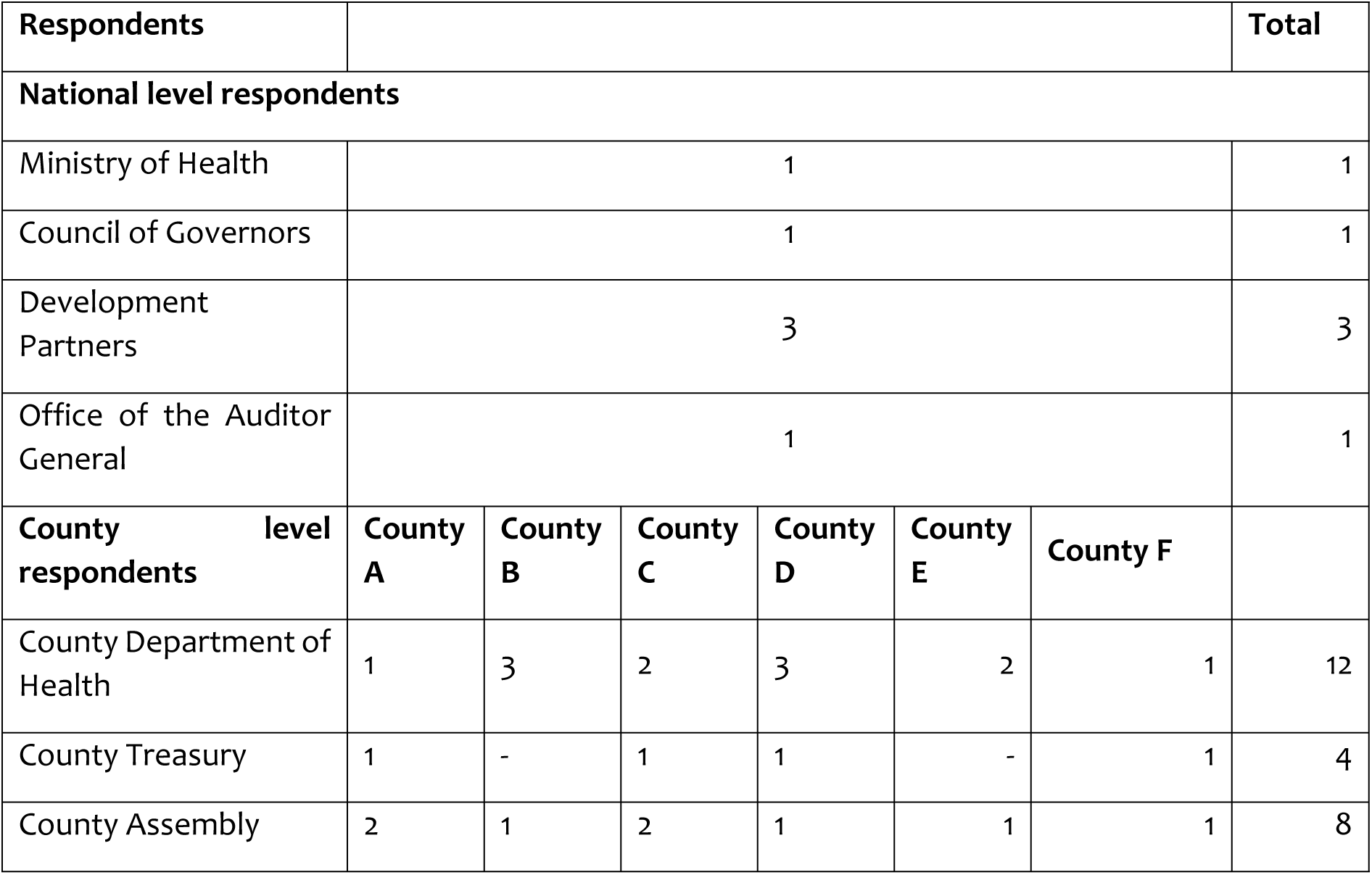
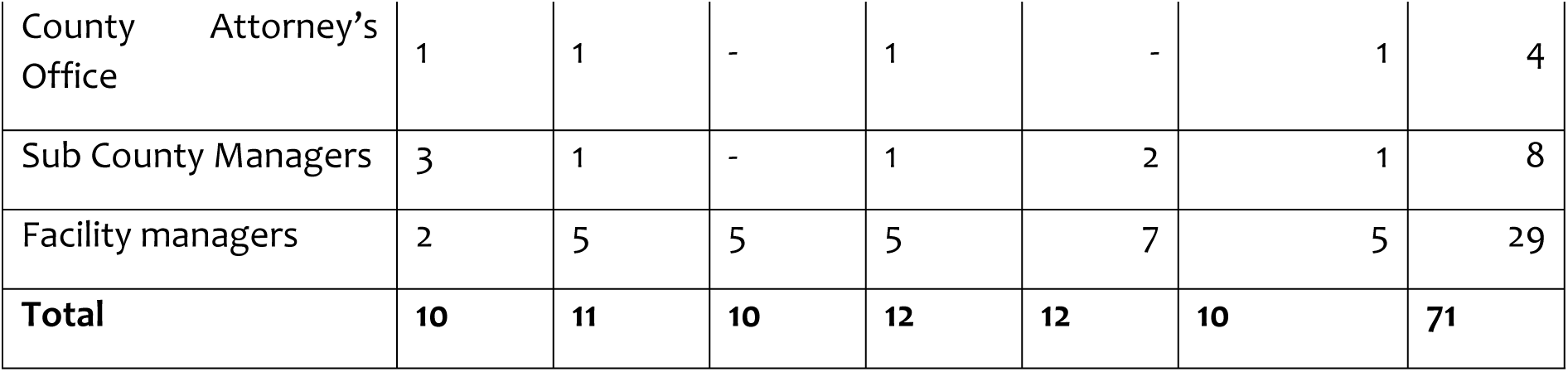
Summary of Study Participants.

| Respondents |  |  |  |  |  |  | Total |
| --- | --- | --- | --- | --- | --- | --- | --- |
| National level respondents |  |  |  |  |  |  |  |
| Ministry of Health | 1 |  |  |  |  |  | 1 |
| Council of Governors | 1 |  |  |  |  |  | 1 |
| Development Partners | 3 |  |  |  |  |  | 3 |
| Office of the Auditor General | 1 |  |  |  |  |  | 1 |
| County level respondents | County A | County B | County C | County D | County E | County F |  |
| County Department of Health | 1 | 3 | 2 | 3 | 2 | 1 | 12 |
| County Treasury | 1 | - | 1 | 1 | - | 1 | 4 |
| County Assembly | 2 | 1 | 2 | 1 | 1 | 1 | 8 |
| County Attorney's Office | 1 | 1 | - | 1 | - | 1 | 4 |
| Sub County Managers | 3 | 1 | - | 1 | 2 | 1 | 8 |
| Facility managers | 2 | 5 | 5 | 5 | 7 | 5 | 29 |
| **Total** | **10** | **11** | **10** | **12** | **12** | **10** | **71** |

#### Quantitative study participants

Within the sampled counties, we selected all county referral hospitals and sub-county hospitals, as across counties, they were most affected by limited autonomy unlike level 2 and 3 facilities that had access to some donor funding. We then proportionately sampled dispensaries and health centers to make up a minimum of 20 facilities per county and a total of 141 facilities across the six counties (Table 3). This study was implemented concurrently, using the same tool with another study investigating the impact of primary care network reforms. Where hospitals were already sampled, we used the data but also sampled a maximum of 20 health facilities per county for the autonomy study, hence for some counties the sample exceeds 20 as the bigger study sampled 444 facilities.

**Table 3:** Study Facilities.

| Facility type | Kisumu | Kajiado | Kakamega | Lamu | Migori | Nakuru | Total |
| --- | --- | --- | --- | --- | --- | --- | --- |
| Dispensary | 4 | 11 | 8 | 26 | 9 | 4 | 62 |
| Health Centers | 4 | 3 | 4 | 3 | 1 | 1 | 16 |
| Hospitals | 16 | 6 | 9 | 3 | 15 | 15 | 63 |
| Total | 24 | 20 | 21 | 32 | 25 | 20 | 142 |

### Data Collection

We provided written informed consent to all study participants. Qualitative interviews were conducted by AM and HO and lasted about 30-60minutes. For the document reviews, we included the national FIF law, the FIF model law, county specific FIF related laws, the PFM Act and proposed FIF regulations. The quantitative health facility assessments on the other hand were conducted by trained data collectors. Data was collected from managers of sampled health facilities using an interview administered structured questionnaire. Pilot assessments were done with non-study facilities in Kisumu, Kiambu and Mombasa counties.

### Data Management and Analysis

We transcribed the audio recordings verbatim into Microsoft Word then imported into NVivo 12 for analysis. We employed a thematic analysis approach, which involved: familiarization with the data, generating initial codes, identifying and reviewing emerging themes, refining and naming these themes, and defining the overarching narrative they conveyed. We held a stakeholder validation meeting in November 2024 to review and confirm the findings, and incorporated feedback from this session into the final analysis.

For the quantitative analysis, we conducted a descriptive analysis of managers’ perceptions of autonomy reforms, the level of autonomy, challenges of autonomy and the unintended consequences of autonomy reforms. We used stata/SE version 18.0 to do the analysis. The qualitative and quantitative data were merged at analysis

## RESULTS

We present the findings as per the conceptual framework for process evaluations. This entails 1) the emergence of autonomy reforms,2) the implementation fidelity 3) implementation experience.

### Emergence of Reforms

The introduction of Facility Improvement Financing (FIF) reforms across countries in Kenya was driven by political, technical, and public demands.

#### Political Drivers

Politically, at the national level, the president included FIF as one of the four laws packaged as his agenda for health reforms in 2023. At the county level both the governors and the members of the county assembly (MCAs) were instrumental in the formulation of the laws, pushed by public demands, technical demands, or pressure from the national government. For example, in County D, health was one key area where the electorate was most dissatisfied with, and when the executive approached the health department for a solution, FIF was raised as an important issue.

*“You know, most of the people’s complaints were going to them (to the MCAs).. Most of the patients going to the hospital and being turned away or missing these things were coming to us for support. Some would even complain that we are not helping them… They’re no, that they’re no that (services). So, we took it as an initiative to find the best solution.” Member of County Assembly, County B*

#### Technical Drivers

Technically, health officials at the county and national levels prioritized health facility autonomy reforms influenced by their knowledge of the effectiveness of FIF reforms both in the pre devolution era and in other counties. Several counties (including all 6 of the selected counties) had already begun initiatives toward similar reforms, prior to the national law, and the Council of Governors and the Ministry of Health (MOH) had drafted a model for facility autonomy reforms, which eventually was adopted to form the FIF law. The technical push was due to the realization that health facility functioning had been compromised by the bureaucratic challenges occasioned by the lack of autonomy. In counties with long standing reforms such as county E, historical knowledge ensured they lobbied for retention of facility autonomy.

*” It was like we are taking resources from finance to health. It was not easy; it was not smooth. There were a lot of hitches and obstacles, but the Department of Health pursued the bill because we had a vision, we knew what FIF could do. By that time, we had a lot of issues like pending bills. Our hospitals were doing badly in terms of commodities. We were not able to procure commodities. We were waiting for the county contribution, and as you know, there are many delays from the National Treasury” County Health Manager County C*

*“…it takes time because we have to make a request (for supplies), the request has to go through those procedures, examination, the internal controls, then you have to request funds from the controller of budget officially. The controller of the budget again takes maybe one week or two weeks. Meanwhile, you have an emergency happening: you don’t have fuel in the hospital, the hospital is temporarily shut down, it’s not running. The turnaround time takes one month for you to get funds…the procedure is the same; you can’t tell the controller of budget it’s an emergency.” County Health Manager County D*

#### Public Needs

There were also public complaints to politicians regarding the dire state of public facilities. This pushed the politicians to investigate the causes of the challenges leading to the identification of facility autonomy as a challenge.

*“… the governor accepted for us to retain the, FIF monies because of the problems that he faced. It was in his first term that health almost costed his governorship…if you look in social media, health is the department that was not doing well. And when he called the department for a meeting, he was told, “Governor if an ambulance breaks, let’s say it is the door that is faulty, you can’t fix it. We told him, “We are ready to change but we need resources in these hospitals.” Casuals used to go on strike because of delayed salaries. So, Governor comes in, the hospital is dirty, he says, “What is wrong?” The hospital says, they don’t have detergents. Detergents are very cheap for a hospital. But, if you don’t have money, you don’t have money and that is when governors saw the sense.” County Health Manager, County C*

##### Implementation Fidelity

###### Policy Design on Paper

Between 2013 and 2022, the MOH and COG issued several circulars and advisories to counties to implement health facility autonomy reforms, but most counties did not fully adopt the recommendations; in the sampled counties, only 3 of the 6 had partially adhered to these circulars. In 2022, the COG developed a model law that was adopted by some counties, including the remaining 3 counties, however they all contextualized the law to their county specific contexts. Finally, in October 2023, the National Government enacted the Facility Improvement Financing (FIF) Act 2023, which was adapted from the model law, and in it allowed counties to form their own legislation to extend the law. Table 4 below shows the key areas in each of the laws – national, COG model, and county specific laws. Five of the six counties enacted their laws before the national law, while one county had started the process of developing the law, but this was enacted after the national law.

**Table 4:**
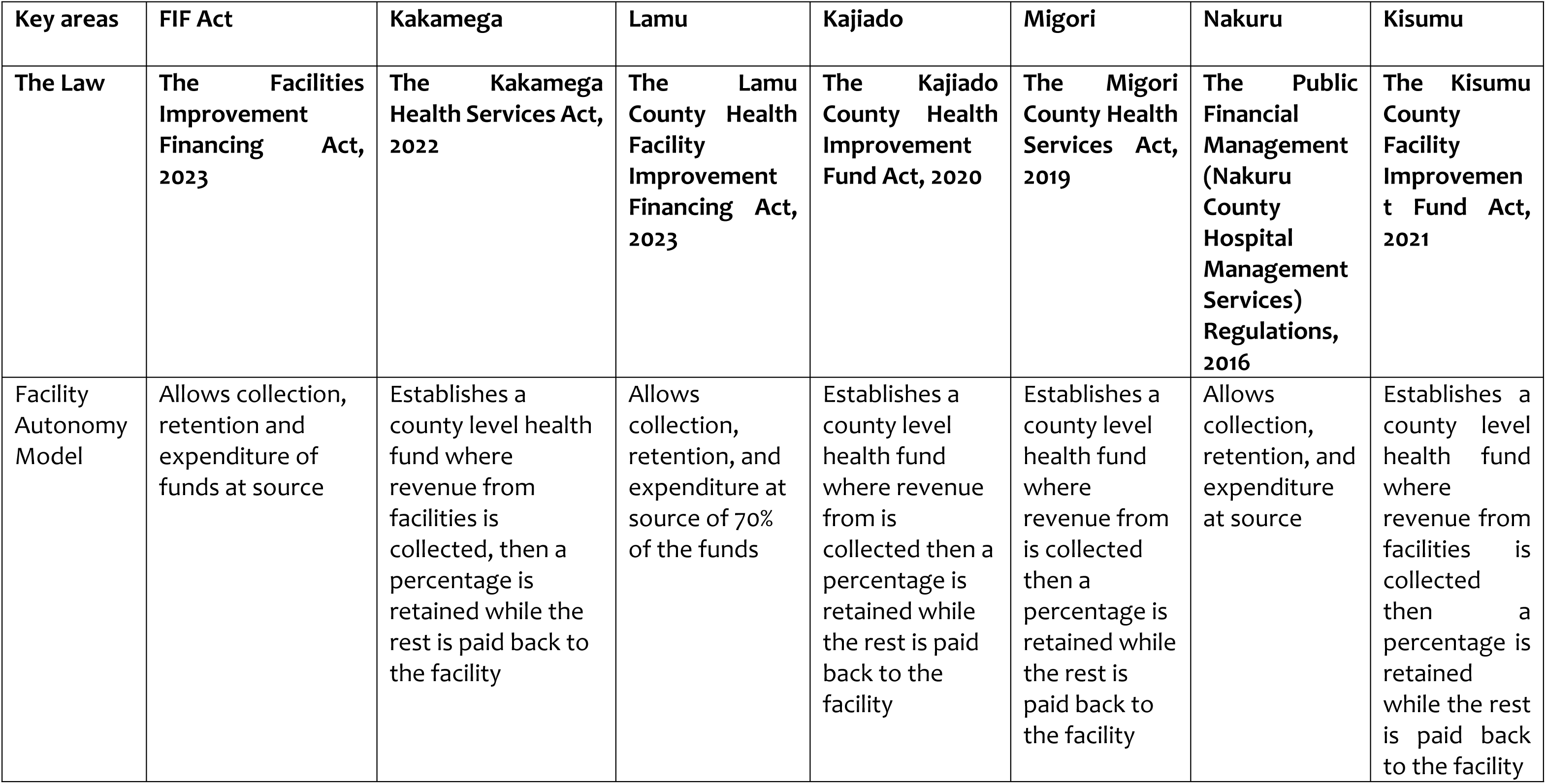
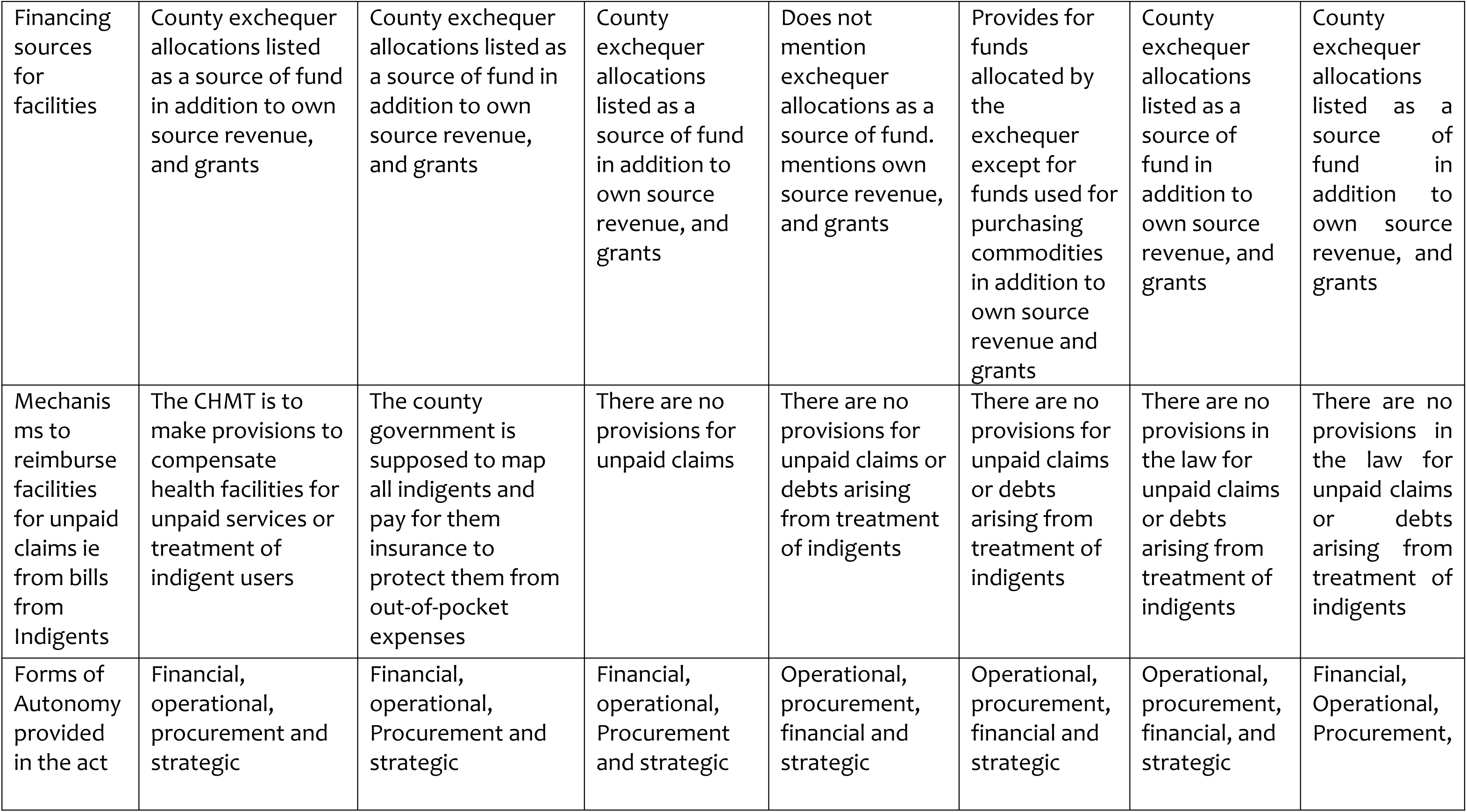
Key areas of concern in FIF laws.

###### Facility financial autonomy model

There were generally two models of autonomy implemented by the counties. One model that was recommended by both the COG and MOH adopted the Appropriation in Aid process guided by Section 109(2)(b) of the PFM act where revenue is collected, retained and spent by the health facilities. Another model, adopted by some counties relied on Section 109(2)(a) of the PFM act and established FIF Fund accounts. In this case, revenue from all facilities were collected and transferred to a central fund account managed by a Fund Manager. In Kajiado, Migori, Kakamega, Kisumu, and Nakuru, funds were collected in a hospital collection account first, then sent to a county health account before part, or all the funds were returned to the hospital. In Kajiado, 70% of the funds were returned to the hospital while 15% was allocated to the County Health Management Team for activities such as supervision, and the remaining 15% was allocated to primary healthcare facilities (level 2 and 3) that did not raise revenue. In Kakamega, 97% was returned to hospitals, while 3% of the funds were used to pay for the funds administration. In Lamu, hospitals generating own source revenue were supposed to retain 70%, and remit 30% to support management functions, primary health care services and community health services. However, since implementation the modalities for implementing this had not been established, the hospitals retained 100% of the collected revenues. In Nakuru, all funds were returned to the hospital accounts. Nakuru county allowed facilities to retain funds at the time of quantitative data collection, however this changed at the time of qualitative data collection, where funds had to go through a special purpose account first before being remitted to facilities.

The retention model involved less bureaucratic processes. However, it was thought to introduce accountability challenges because the funds were easily accessible and lacked the accountability mechanisms of the Public Finance Management system such as the digital tracking of expenditure and approvals. While there were no reported events of the facilities misusing funds some counties reported that sometimes the county treasury and the governor’s office would request for funds from the facility for non-health related activities. In some instances, these money would be refunded, while in others, they were not.

*“Then the other issue is, within health department, there are interests coming up within the department. We (health system actors) have fought for 11 years, we have gotten the money away from CRF [County revenue Fund], now it’s our time to have the money that we have now within our department. We are fighting for autonomy so, that the money goes to the facility to improve services and not at the department to be used for other things. So, yeah. Just to make sure that we don’t lose sight of the focus that ideally FIF the idea behind it is improvement of services. “ National Government Actor*

The fund model was considered more efficient than the previous County Revenue Fund (CRF) by facilities, but it was still viewed as an additional layer of bureaucracy. It was perceived to introduce a parallel management structure with additional administrative costs. Additionally, it did not allow health facilities to retain and use all the money that they raised - in many cases, facilities got only a fraction of what they collected. The reasons for the creation of the fund model varied. In some instances, it was created to satisfy political interests, in others to provide accountability, and in others to provide for funding to other CDoH entities like the CHMT, Sub County Health Management Team (SCHMT) and lower level (PHC) facilities that did not charge user fees.

*“I think the only deviation I see was that the point where now, the monies then had to be channeled into a pooled or rather another fund which I call a mini CRF and its purpose was not so clear” Sub County Manager County B*

*“… For us we told you they retain 70 we take 30 because of the health emergencies, primary health care and now supporting the CHMT so if they retain and I know you have visited counties, and you know accessing funds from county treasury is so hard…. The question is … if the hospitals are empowered and the CHMT are not empowered, is that department going to work? So the Waziri (County Executive Committee Member) would be like the med sup of the hospital…” County health Manager County C*

###### Mechanisms for reimbursing facilities for non-payment such as by indigent users

The national FIF law requires the CHMT to make provisions to reimburse facilities for services provided to indigents. Kakamega’s law provided mechanisms to cover indigents from financial difficulties by registering them with the social health insurance. Kajiado’s, Kisumu’s, Lamu’s, Migori’s, and Nakuru’s FIF laws have no provisions for indigents.

###### Types of autonomy granted

All the laws, county and national, convey to facilities some form of financial autonomy, for some to collect, retain and spend, others to collect, remit to a fund, receive funds back and spend. The laws also convey some form of procurement autonomy, although no limitations are provided in the FIF law, these are interpreted together with the Public Procurement and Asset Disposal Act. One county, Migori County limited facilities from the purchase of health commodities and retained it as a function of the county. All the laws also convey autonomy over their strategic plans, and operations, through functions of the health facility management boards and committees where they are mandated to make their plans and budgets and to make decisions on facility operations.

###### Deviation between Policy on Paper and Policy in Practice (In) Fidelity

The first deviation occurred when county treasuries either failed to provide complementary funding or reduced it significantly. County treasuries expected facilities to rely solely on revenue the facilities generated, even though the national and county specific laws expected them to supplement existing funding. The agreement to supplement was from the consensus that own source revenue on its own was insufficient to sustain facility operations. For example, in county A, their law required monthly transfers from allocated funds from the exchequer to the facilities. However, this was not done. The treasury’s resistance originated from their initial opposition to the policy reforms, as it required them to relinquish some control. Consequently, they leveraged their authority to withhold additional resources from the health department.

*“This journey towards independence in health financing has met a lot of resistance from the treasury. The money(to complement) is appropriated, it is budgeted for, but now the treasury feels that the money the hospitals are generating should be sufficient to facilitate all the activities but it’s not the position.” County Assembly County A*

The second deviation occurred in the CHMT’s duty to make provisions to reimburse facilities for services rendered to indigent patients or those unable to pay as per the national FIF act. In practice, facilities in all counties that have implemented the reforms bore the full cost of these waivers and exemptions themselves. In other counties, waivers were banned and considered unacceptable – patients had to bear the full cost, but if the case really required waivers, then a maximum waiver of 25% would be given (County F). For counties where interviews were conducted post October 2024 (when Social Health Insurance reforms were rolled out), the social insurance mitigates the need for waivers as the insurance cover did not have a waiting period (County E and F).

*“Unfortunately, that is not compensated for either by the government or by any other partner or well-wisher so it’s a loss,and this has grossly affected level five which is the referral facility. Most of those referral emergencies, very critical cases, end up in ICU and with no one to facilitate, so the facility ends up using resources that are never reimbursed. So they end up just making losses in terms of waiver.*.” Sub County manager County A

The third deviation was on the level of autonomy granted, while the laws on paper granted financial autonomy, only 59% reported autonomy to spend social insurance payments, 51% county funds and 77% autonomy to spend user fees. 95% reported autonomy over spending priorities (Table 5). Quantitatively, for counties that allowed facilities to retain their funds, autonomy over county funds was 15% for Nakuru and 24% for Migori (Table 5). For facilities that first send funds to a county level fund account then the autonomy over county funds was perceived to be higher, Kisumu (71%), Kajiado (63%) and Kakamega (81%). Across the counties level 2 and 3 facilities did not collect user fees, so they received allocations from the exchequer and therefore had some form of autonomy over exchequer allocations, level 4 and 5s were expected to fully manage all recurrent costs except salaries, hence had no autonomy over exchequer allocations, despite having allocations, spending decisions were made centrally.

**Table 5:** Percentage of Facilities Reporting Financial Autonomy.

| subgroup | Spending Priorities | County Funds | Danida Funds | Procurement Autonomy | NHIF Funds | User Fees |
| --- | --- | --- | --- | --- | --- | --- |
| **By county** |  |  |  |  |  |  |
| Kajiado | 94% | 67% | 100% | 78% | 67% | 50% |
| Kakamega | 100% | 69% | 60% | 92% | 62% | 75% |
| Kisumu | 100% | 74% | 86% | 87% | 83% | 94% |
| Lamu | 92% | 77% | 69% | 92% | 23% | 0% |
| Migori | 96% | 24% | 80% | 76% | 48% | 67% |
| Nakuru | 84% | 16% | 50% | 89% | 63% | 80% |
| **By level of health facility** |  |  |  |  |  |  |
| Dispensary | 90% | 65% | 78% | 85% | 38% |  |
| Health Center | 100% | 45% | 82% | 64% | 45% |  |
| Hospital | 97% | 43% |  | 88% | 77% | 77% |
| Total | 95% | 51% | 80% | 85% | 59% | 77% |

While the laws granted some level of procurement autonomy, only 85% of the facilities had autonomy over what to procure (Table 5) , 39% had autonomy over the choice of suppliers and 82% had autonomy over the quantities to be procured (Table 6). In some counties, the level of procurement autonomy was limited by availability of procurement personnel – the higher-level facilities were allocated procurement personnel while the lower levels were not. In other counties, facilities were prohibited from spending on certain items - County D prohibited procurement of medicines and supplies On the choice of suppliers, in some counties, all facilities were limited to prequalified suppliers (County C, F), while in others (County B, A) higher level facilities could tender but lower levels were limited to prequalified suppliers.

**Table 6:** Percentage Facilities with Procurement Autonomy.

|  | Choice of Quantities |  | Choice of Suppliers |  | Choice of Commodities |  |
| --- | --- | --- | --- | --- | --- | --- |
| subgroup | On paper | In practice | On paper | In practice | On paper | In practice |
| **By county** |  |  |  |  |  |  |
| Kajiado | 78% | 78% | 44% | 44% | 78% | 78% |
| Kakamega | 92% | 92% | 46% | 46% | 92% | 92% |
| Kisumu | 100% | 91% | 70% | 70% | 96% | 87% |
| Lamu | 85% | 85% | 8% | 8% | 92% | 92% |
| Migori | 68% | 68% | 16% | 16% | 76% | 76% |
| Nakuru | 89% | 84% | 42% | 42% | 95% | 89% |
| **By level of facility** |  |  |  |  |  |  |
| Dispensary | 72% | 70% | 17% | 17% | 88% | 85% |
| Health Center | 91% | 73% | 18% | 18% | 82% | 64% |
| Hospital | 92% | 92% | 57% | 57% | 88% | 88% |
| Total | 85% | 82% | 39% | 39% | 87% | 85% |

*“In XXX procurement is done at three levels, we have the county level that deals majorly with the big projects and equipment. We have at the Sub County level, XXX County Referral Hospital, generates a list of pre-qualified suppliers who are able to supply the facilities at the Sub County levels…What is done is that because of shortage of procurement officers, the procurement of these services are done at the XXX County Referral Hospital level…the facility will then sit down see who is best suited to supply them (from prequalified list).” Sub County Manager County B*

None of the FIF laws directly granted human resource autonomy. The general understanding across counties was that human resource recruitment and management is a preserve of the county public service board (CPSB). Nonetheless, financial autonomy allowed facilities to recruit support staff, with 66% of facilities reporting they had the authority to do so (Table 7). However, in some counties, after the facilities had this autonomy for years, the CPSB retracted this mandate (County B, E). Only 11% of facilities reported having autonomy to recruit clinical staff (Table 7). This limitation was due to various factors, including legal restrictions in certain counties, such as County B, and E, where the recruitment of staff was under the mandate of the CPSB. Additionally, in other facilities, the revenue generated was insufficient to cover the costs of recruiting clinical staff.

**Table 7.** Percentage Facilities with Human Resource Autonomy.

| subgroup | Authority to discipline staff | Authority to fire staff | Authority to recruit clinical staff | Authority to recruit support staff |
| --- | --- | --- | --- | --- |
| **By county** |  |  |  |  |
| Kajiado | 56% | 56% | 11% | 89% |
| Kakamega | 62% | 46% | 15% | 62% |
| Kisumu | 91% | 78% | 13% | 100% |
| Lamu | 46% | 8% | 0% | 23% |
| Migori | 60% | 32% | 4% | 76% |
| Nakuru | 79% | 21% | 16% | 26% |
| **By level of health facility** |  |  |  |  |
| Dispensary | 40% | 25% | 7% | 62% |
| Health Center | 73% | 55% | 18% | 91% |
| Hospital | 85% | 52% | 10% | 65% |
| Total | 68% | 42% | 10% | 67% |

###### FIF reform implementation

In our theory of change, we listed activities based on literature and stakeholder feedback that are required to optimize health facility autonomy reforms (15). An evaluation of implementation on the ground showed that most activities were either partially implemented or not implemented (Table 8). Only three issues were fully implemented; accountability mechanisms – supervision and reporting, aspects of financial autonomy – related to opening of bank accounts, and third, strategic autonomy-aspects related to development of annual work plans. These were mostly activities that were already ongoing and were only extended to accommodate financial autonomy reforms.

**Table 8:**
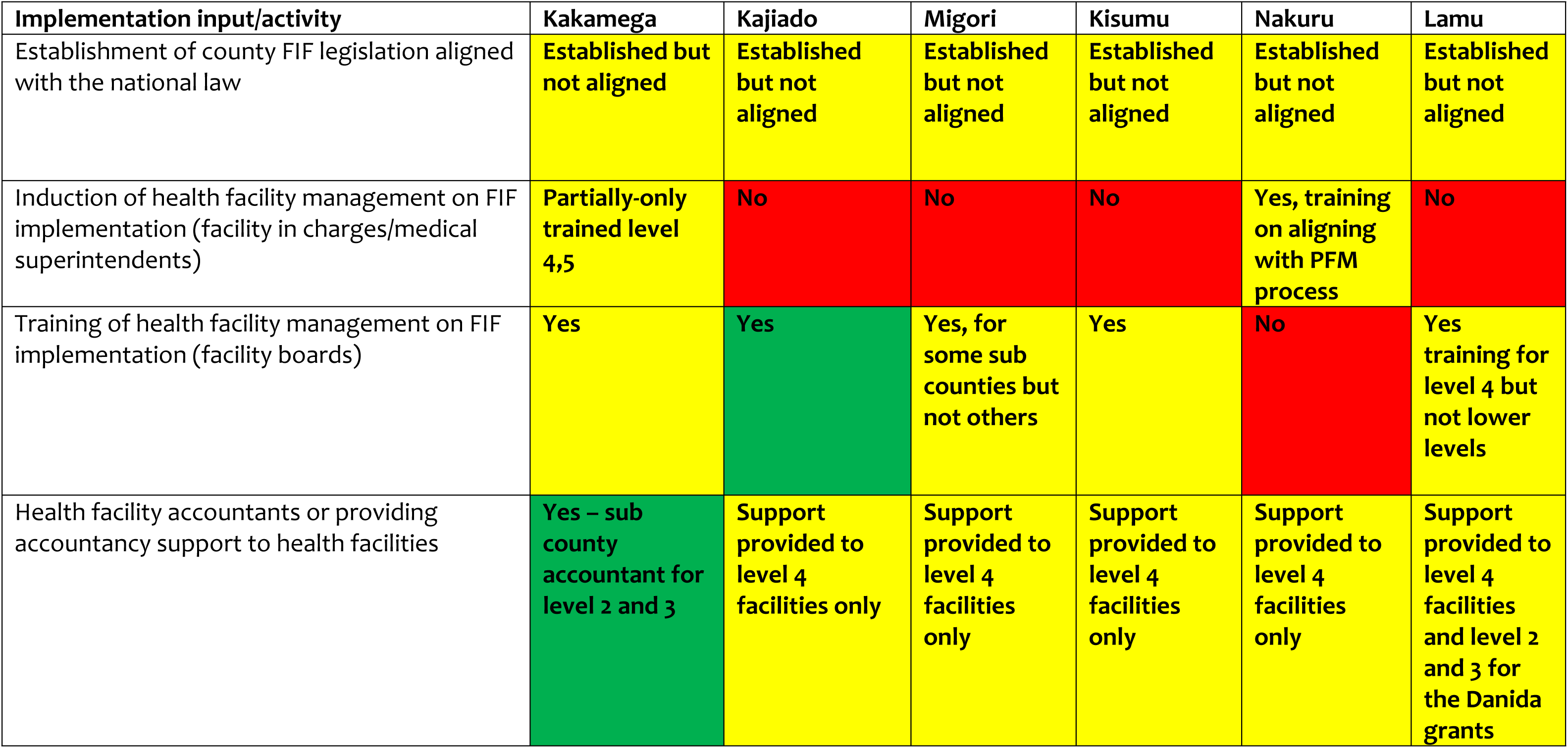
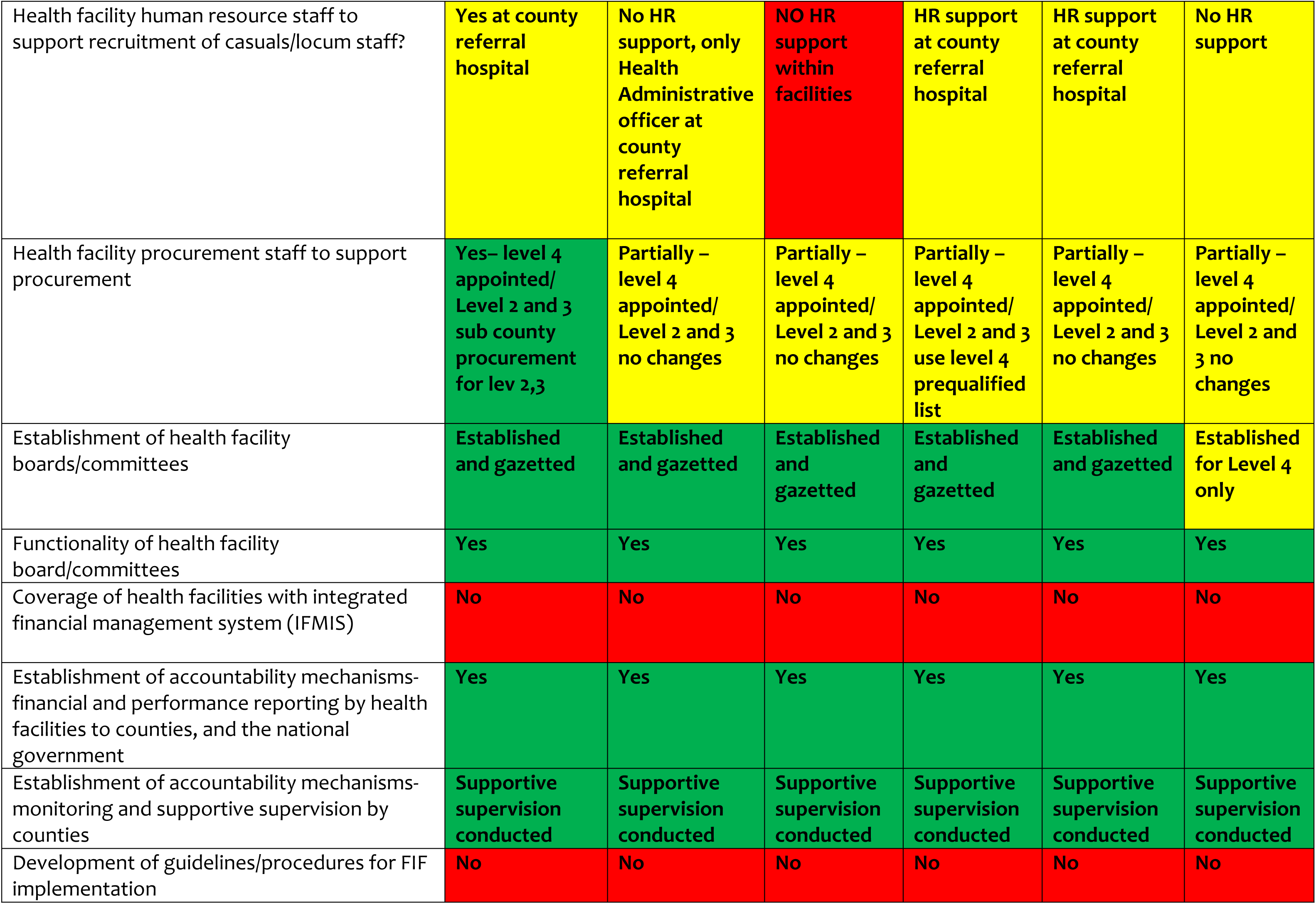
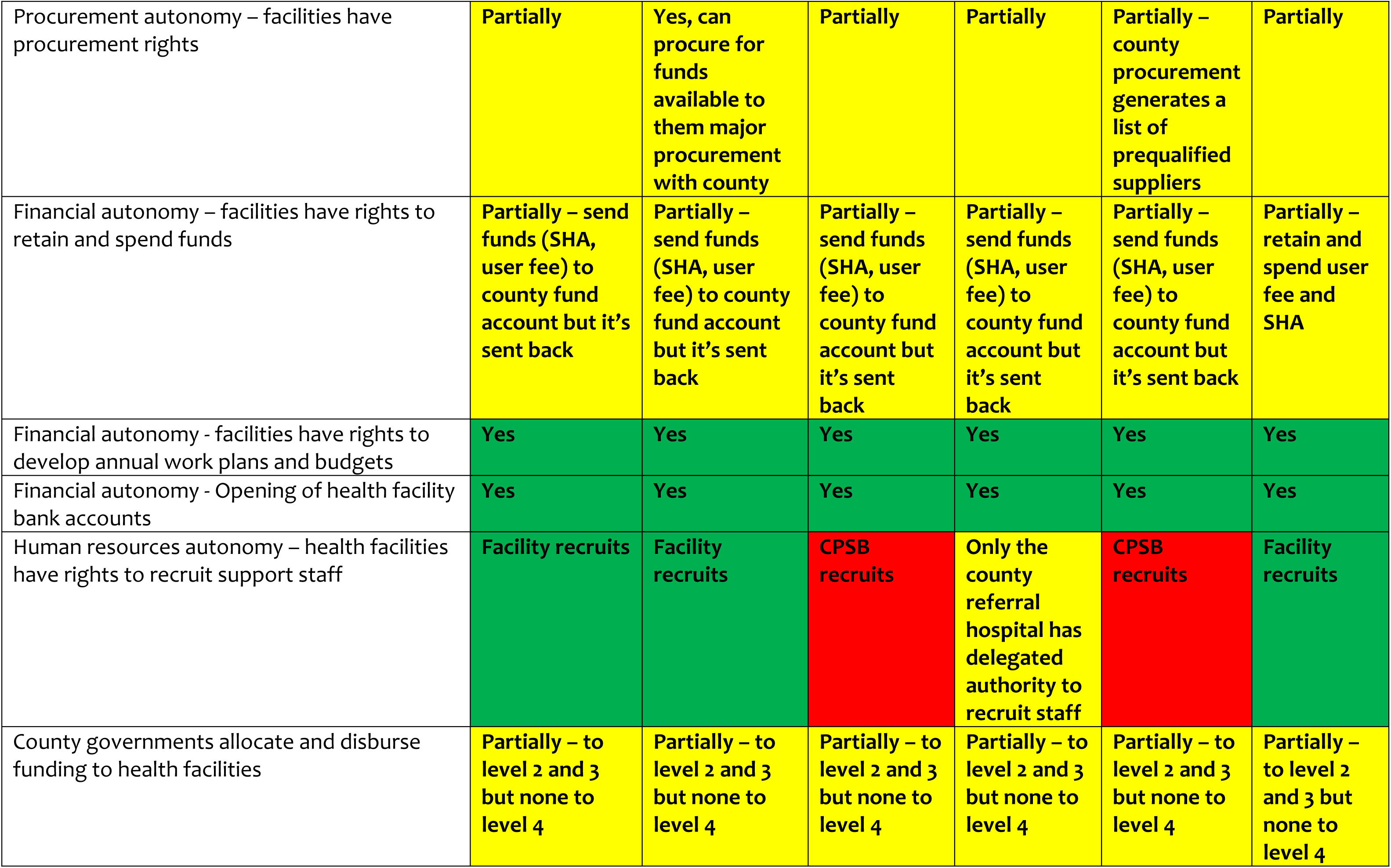
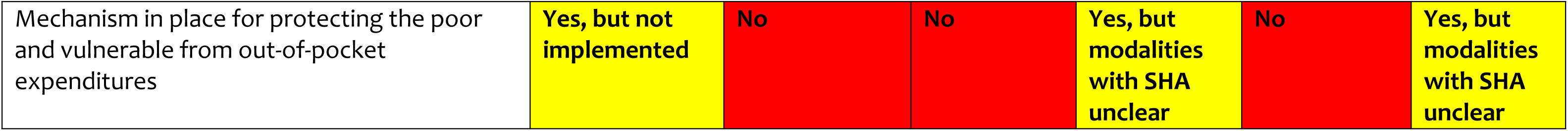
Activities and Inputs to the reforms.

| Implementation input/activity | Kakamega | Kajiado | Migori | Kisumu | Nakuru | Lamu |
| --- | --- | --- | --- | --- | --- | --- |
| Establishment of county FIF legislation aligned with the national law | Established but not aligned | Established but not aligned | Established but not aligned | Established but not aligned | Established but not aligned | Established but not aligned |
| Induction of health facility management on FIF implementation (facility in charges/medical superintendents) | Partially-only trained level 4,5 | No | No | No | Yes, training on aligning with PFM process | No |
| Training of health facility management on FIF implementation (facility boards) | Yes | Yes | Yes, for some sub counties but not others | Yes | No | Yes training for level 4 but not lower levels |
| Health facility accountants or providing accountancy support to health facilities | Yes – sub county accountant for level 2 and 3 | Support provided to level 4 facilities only | Support provided to level 4 facilities only | Support provided to level 4 facilities only | Support provided to level 4 facilities only | Support provided to level 4 facilities and level 2 and 3 for the Danida grants |
| Health facility human resource staff to support recruitment of casuals/locum staff? | Yes at county referral hospital | No HR support, only Health Administrative officer at county referral hospital | NO HR support within facilities | HR support at county referral hospital | HR support at county referral hospital | No HR support |
| Health facility procurement staff to support procurement | Yes– level 4 appointed/ Level 2 and 3 sub county procurement for lev 2,3 | Partially – level 4 appointed/ Level 2 and 3 no changes | Partially – level 4 appointed/ Level 2 and 3 no changes | Partially – level 4 appointed/ Level 2 and 3 use level 4 prequalified list | Partially – level 4 appointed/ Level 2 and 3 no changes | Partially – level 4 appointed/ Level 2 and 3 no changes |
| Establishment of health facility boards/committees | Established and gazetted | Established and gazetted | Established and gazetted | Established and gazetted | Established and gazetted | Established for Level 4 only |
| Functionality of health facility board/committees | Yes | Yes | Yes | Yes | Yes | Yes |
| Coverage of health facilities with integrated financial management system (IFMIS) | No | No | No | No | No | No |
| Establishment of accountability mechanisms- financial and performance reporting by health facilities to counties, and the national government | Yes | Yes | Yes | Yes | Yes | Yes |
| Establishment of accountability mechanisms- monitoring and supportive supervision by counties | Supportive supervision conducted | Supportive supervision conducted | Supportive supervision conducted | Supportive supervision conducted | Supportive supervision conducted | Supportive supervision conducted |
| Development of guidelines/procedures for FIF implementation | No | No | No | No | No | No |
| Procurement autonomy – facilities have procurement rights | **Partially** | **Yes, can procure for funds available to them major procurement with county** | **Partially** | **Partially** | **Partially – county procurement generates a list of prequalified suppliers** | **Partially** |
| Financial autonomy – facilities have rights to retain and spend funds | **Partially – send funds (SHA, user fee) to county fund account but it's sent back** | **Partially – send funds (SHA, user fee) to county fund account but it's sent back** | **Partially – send funds (SHA, user fee) to county fund account but it's sent back** | **Partially – send funds (SHA, user fee) to county fund account but it's sent back** | **Partially – send funds (SHA, user fee) to county fund account but it's sent back** | **Partially – retain and spend user fee and SHA** |
| Financial autonomy - facilities have rights to develop annual work plans and budgets | **Yes** | **Yes** | **Yes** | **Yes** | **Yes** | **Yes** |
| Financial autonomy - Opening of health facility bank accounts | **Yes** | **Yes** | **Yes** | **Yes** | **Yes** | **Yes** |
| Human resources autonomy – health facilities have rights to recruit support staff | **Facility recruits** | **Facility recruits** | **CPSB recruits** | **Only the county referral hospital has delegated authority to recruit staff** | **CPSB recruits** | **Facility recruits** |
| County governments allocate and disburse funding to health facilities | **Partially – to level 2 and 3 but none to level 4** | **Partially – to level 2 and 3 but none to level 4** | **Partially – to level 2 and 3 but none to level 4** | **Partially – to level 2 and 3 but none to level 4** | **Partially – to level 2 and 3 but none to level 4** | **Partially – to level 2 and 3 but none to level 4** |
| Mechanism in place for protecting the poor and vulnerable from out-of-pocket expenditures | Yes, but not implemented | No | No | Yes, but modalities with SHA unclear | No | Yes, but modalities with SHA unclear |

###### Implementation of Health Facility Autonomy Reforms

##### Implementation experience

###### Enablers of Implementation

During the implementation, there were many enablers of FIF implementation, the most significant were political goodwill, support from development partners, institutional memory and technical capacity of the department of health, consensus among stakeholders and support from county treasury.

Counties with strong political support found it easier to implement autonomy reforms, primarily due to the support from the executive and the legislature (county assemblies) assemblies. Governors who were in favor of these reforms incorporated FIF into their manifestos and issued directives to ensure their implementation. In contrast, governors opposed to the reforms instructed their county treasuries to halt progress. For instance, in Counties A and D, although FIF laws had been passed under previous administrations, they were only enacted when new governors, committed to the reforms, took office.

*“ I had a chat with the ministers of finance. They say they were told not to allow health facilities to retain the money. Yes. Instructions from the governor. Then the governor goes to the health facility and finds that there are no drugs, it’s dirty. And then on the spot demotes, the medical superintendent, what did he expect them to do? And they had not received money the whole year… But the reality is they are acknowledging there’s a problem, but they’re not willing to do what is required to fix it, yet it is within their power to fix it.” National Development Partner*

Although political support was crucial, it largely depended on guidance from technocrats. Those with institutional memory from before the devolution of health services were particularly well-positioned to drive the reforms, as their prior knowledge helped them understand what worked and how. This institutional memory was critical within both the health departments and county treasuries. For example, in County C, health technocrats effectively communicated to politicians the specific challenges that had led to poor health system performance, narrowing down to limited autonomy. In County B, technocrats managed to sustain the reforms during the transition from national to county governments in 2013, but these were later rolled back when other changes, such as the introduction of the UHC pilot in 2019, which offered free healthcare for all, were implemented. This shift limited the autonomy of the system, as autonomy reforms were pegged on revenue collection, but this was

*“Each county had its challenges. Fortunately we didn’t have a challenge because we had a very responsible Assembly, a very responsible Government, and then you know we had been inherited from the provincial administration…we had quite some knowledge and experiences that was very useful to XXX County then..” County Manager County B*

*” if you also have technical people who have institutional memory at the treasury, obviously things will work and besides that the most important person is the governor. You know government works in such a way that, if the government or the head of the government gets the right advice things will work well but if they get wrong advice then that is when you find that other county’s things will change” County Manager county A*

Support from the county treasury was also considered crucial as they provided the necessary systems to successfully establish payment and accountability systems for facility funds. Some counties assigned accounting and procurement staff to their health facilities, facilitating the FIF process and enabling financial and procurement autonomy.

*“we have the facility in charge, we have the administrator, then in each facility there is an accountant, there is a procurement person, so, they jointly work to ensure that accountability done. Yeah.” Sub County Manager County A*

Consensus among stakeholders also played a crucial role in the reforms forward.

***“****autonomy works **if** there’s support from all stakeholders, the hospital, the department, the executive, and also the assembly, and also the facility itself” Member of County Assembly, County C*

*“The county treasury, county assembly, and the department of health, that axis needs to speak in harmony. At the national level, the national treasury… A treasury is an enabling as far as the custodian of PFM, they’re supposed to deploy it in a way that is an enabling, not a barrier, to the other departments. So, the way county treasury players understand this is important…” Development Partner*

Development partners also made it easy to roll out the reforms by providing technical expertise to first, advocate for formulation of the laws and to draft the laws. In some counties such as county D, they provided the resources to train facility boards and committees.

###### Challenges of reforms

Counties that have implemented the reforms have encountered numerous challenges, including resistance from the county treasury, limited capacity to formulate FIF laws, and limited facility capacity to provide services and therefore raise revenue.

County treasury resistance was particularly pronounced in counties where the health department’s own source revenue constituted a significant portion of the county’s revenue. The county treasuries viewed allowing the health sector to retain these funds as a loss of control over the resources. Additionally, the treasury was concerned about the health department’s ability to effectively account for the money. Consequently, they were reluctant to cede power and resources to the health sector.

*“Well, the journey to health financing has been daunting. There has been a lot of resistance because health is a major income generating facility in any County Government. With the fight towards autonomy in financing in the health facilities the treasury felt as if some of their powers are being clipped off and that is why we experienced a lot of resistance despite the fact that the Public Finance Management Act allows for ring-fencing of funds under certain circumstances.” County Attorney, County A*

*“…Somebody has to account for that money. It is not free money. Before we had challenges of accountability that you’d have the hospital wanting to spend it the way it wishes to. And it is not correct. Any public funds has to be appropriated, expensed according to law and accounted for. “County Finance Manager County C*

Regarding implementation capacity, some counties initially enacted laws that were later rejected by the office of the Controller of Budget - a constitutional institution that oversees the implementation of government budgets including the withdrawal of public funds from government accounts. The laws were rejected because they were termed regulations that had a maximum 10-year life span. This rejection led to a cycle of repeatedly drafting and revising these laws. The county technocrats lacked the expertise to effectively formulate the laws, resulting in years of ongoing revisions and repeals.

*” we didn’t have the right expertise… first of all we did it as a regulation and a regulation can be done away with anytime it is not a law. when it went to the controller of budget then they were not okay with some sections of that law. So, when we sought the advice from those other institutions like controller of budget and some of us came on board and we saw the sense of why it was supposed to be a law, we incorporated it in our law” county health manager county A*

Facilities still face challenges with revenue generation, largely due to their poor prior conditions. Some facilities were almost totally dysfunctional, and did not provide diagnostics, and procedures that would generate revenue. Counties were expected to equip and operatiomalize these facilities before introducing autonomy reforms, but this was not fully achieved. As a result, many facilities are unable to offer a comprehensive range of services, limiting their potential to raise revenue.

###### Outcomes of reforms

Study participants reported that facility autonomy reforms have had several positive effects on health facility functioning and performance, including enhancing decision making, human resources for health, facility revenue collection, procurement and supply chain and infrastructural improvements.

The respondents noted that facility autonomy reforms have simplified decision-making processes, enabling facility managers to respond promptly to emergencies within health facilities by providing flexible funding that can be allocated according to their immediate needs. According to the managers, this has protected facilities from the bureaucratic obstacles that have historically affected access to government exchequer funds.

*“Yes, they are helpful. They help cater to emergency needs that arise within the county or hospitals. Because of the periodic nature of how funds are received by the county government, FIF is able to meet those gaps in between.” County Finance Manager County C*

*” just knowing that I’m in the control seat, I don’t have to rely on someone else. Before you would have to do lots of letters. Then those letters can even go and just sit on someone’s desk forever. You’re there waiting and waiting and waiting for a response. For instance, an essential commodity, like oxygen. I’m relying on cylinders to supply oxygen. Once mine have been depleted, if plans have not been made properly at the county level, then it means I have to keep on asking oxygen, oxygen, oxygen, and as long as someone else has to approve before I can get that oxygen, there’s that patient who’s likely to die waiting…” Facility Manager County A*

FIF reforms have facilitated the timely payment of various service providers such as locum staff and suppliers, thereby enhancing the availability of services. With direct access to funding, facilities were able to process payments more efficiently.

*“…casuals are paid they never used to be paid for several months.” County Health Manager County B*

These reforms have also enhanced staff satisfaction by providing a conducive work environment equipped with the necessary tools for effective service delivery and by increasing the overall staff count.

*” Now they have whatever it is they need, they have that a conducive environment. before you’d have challenges like I don’t have gloves how am I supposed to touch a patient without gloves, now these are available… So the work environment is much, much better….” Facility Manager County A*

The respondents noted that autonomy reforms increased revenue collections and strengthened accountability systems for managing these funds. Providers have become more diligent in billing patients and ensuring that all revenue is properly collected as they now play a more active role in the decision making and they are certain of what the funds they collect are used for. Furthermore, the increased availability of services has attracted more patients to public health facilities, hence increasing revenue. For instance, in County C, the CHMT observed improved availability of medicines contributed to higher patient volumes.

*“It creates responsibility, it makes the people who are involved in the collection of this money, really own the process. You can imagine that you collect money, you take it to some place and it doesn’t come back, what energy do you have to collect that money? The energy that you would have then is to steal the money, because in any case somebody is using the money elsewhere…. You might as well steal it, you know. So, the fact that our facilities receive their money encourages people to do more.” County Health Manager County B*

Autonomy reforms were linked to more timely procurement in addition to increased availability of health products and technologies. With greater autonomy, facilities have been able to establish closer, more personalized relationships with suppliers, allowing them to place orders on credit and guarantee payments. In some counties, suppliers have more confidence in dealing directly with the facilities than with the county government.

*” I think the delays in supplies has really gone down. If you talk to the common mwananchi, they’ll tell you that things are moving in the facility. majorly its because we engage with our suppliers and we are able to pay them for the services and that wouldn’t have been possible if we didn’t have FIF because at least I know what it is I’ve been able to collect and I know I’ll be able to pay this and that supplier*.” Facility Manager County A

Health facility autonomy reforms have allowed the renovation and construction of facilities to ensure that facilities can adequately provide for client needs. Because facilities have direct access to funds, they can make priorities and therefore prioritize things are significant for the running of facilities such as repairs and maintenance.

*“Compared to 4 years ago, right now if you go the facility you will notice a big difference. People are able to maintain the facility, they can paint it” County Health Manager County B*

Unintended consequences of the reforms

There are several unintended consequences that emerged due to autonomy reforms including: defunded primary healthcare facilities, barriers to access for the poor, increased cost of services, increased focus on revenue generating schemes, and misappropriation of funds. These unintended consequences stemmed from counties’ deviations from the intended implementation, or the failure to anticipate some of the documented repercussions, and therefore failure to make provisions in policy to mitigate the unintended consequences. The deviations in implementation compromised the mechanisms the laws had put in place to protect both facilities and citizens from the unintended consequences of health facility autonomy.

First, there was limited complementary funding from the exchequer as identified in the deviations. While the limited funding affected all facilities, lower-level facilities that did not collect user fees were worse off, unfunded and therefore dysfunctional. 36% of the facility managers either strongly agreed or agreed that autonomy reforms have resulted in dependence on user fees. This was higher among counties that had implemented autonomy reforms for longer, like Nakuru where 63% of the facility managers either strongly agreed or agreed that autonomy reforms resulted in dependence on user fees.

*“The fact that it’s like funding for primary health care is not really being implemented. It’s not being implemented at all. Leaving now the lower-level facilities at the mercies of their level 4 facilities to support them when they run out of commodities. ” Sub County Manager County A*

*“we are moving although we are not yet there, we are moving towards the right direction and we’ve been able now to buy commodities….we recently did a baseline survey in our health facilities when we were doing primary care networking in XXX and the finding was that in level 4 facilities where they collect the FIF, the commodities were constantly almost always there as opposed to the level 2 and 3 facilities that where we had stock outs for major parts of the year.” Sub County Manager County B*

Second, counties have yet to establish effective mechanisms of reimbursing facilities for providing healthcare to patients who can’t afford to pay. Facilities are expected to bear the cost of providing these services. As a result, providers who currently bear the cost of treating indigents, are becoming hesitant about providing services to the poor, who cannot afford user fees, with a potential of this escalating to denied services. In some counties, they require patients to pay whatever they can before they are waivered. In fig 1 below, more than 30% of providers in Nakuru, Migori and Kisumu counties believe that facility autonomy reforms have resulted in increased focus on revenue over provision of basic services.

**Fig 1:**
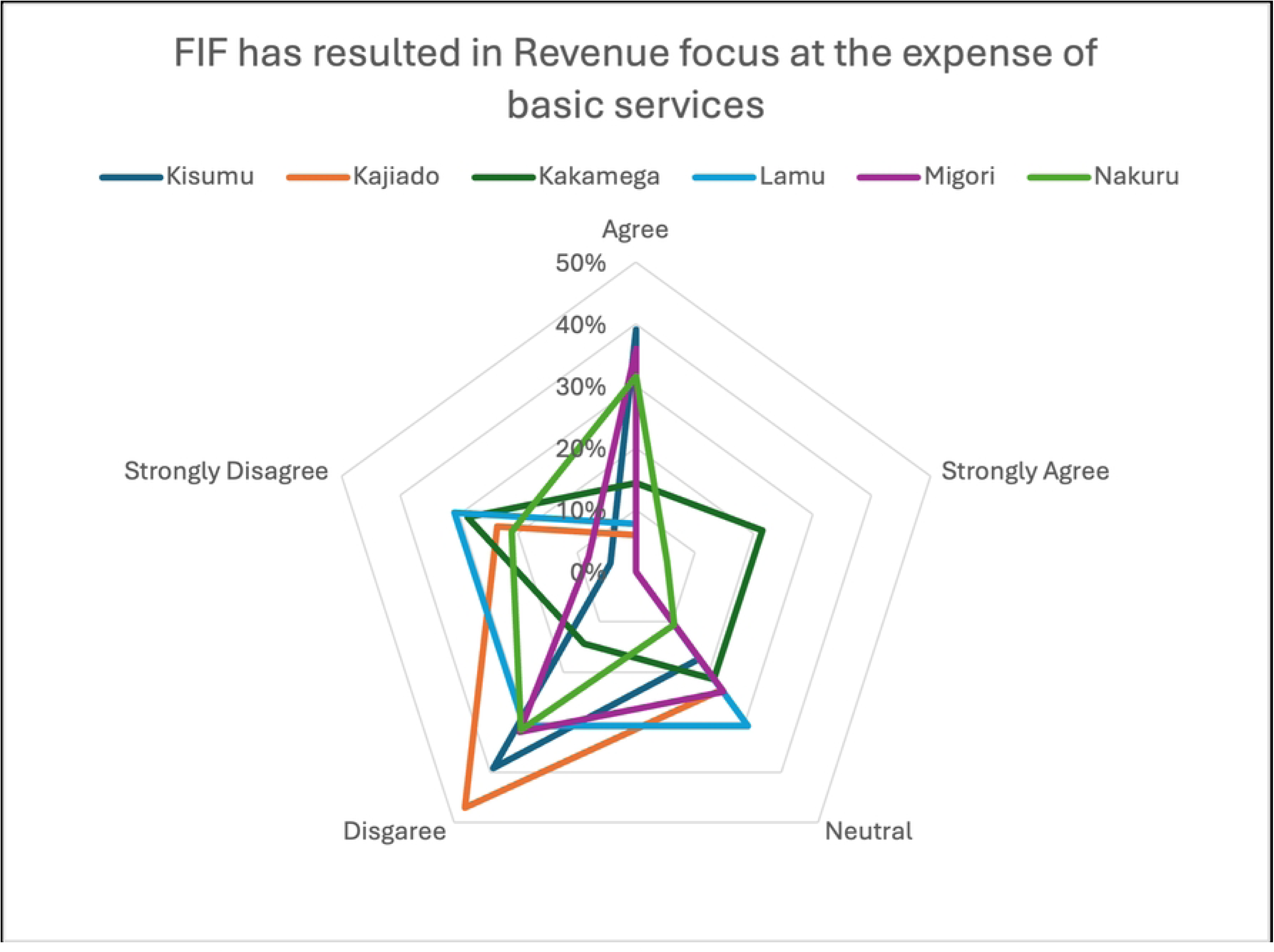
Facility Managers Perception on FIF resulting in a focus on revenue generation over provision of basic services.

*“the county has tried to come up with a waiver and exemptions policy we should be sitting sometime this week to just ratify it …I think the only thing that needs to come out clear and I’m hoping that it has now*

*been captured is exactly who pays for the service that has been waived and exempted. Because as long as it’s still silent on that I foresee a time when we will not be able to continue offering that service if someone doesn’t really come out clearly….” Facility Manager County A*

*“Well as I’ve said before XXX people don’t have money as such. The larger population don’t have money as compared to the other regions our neighbors here XXX, …. We work to eat and live so by bringing this FIF Act it has led to an increase of waiver in our facilities as I said before and an increase of waiver means we are decreasing the access because the patient can’t afford.” Sub County Manager County B*

Third, the respondents expressed fears that facilities will focus on revenue generating services at the expense of basic services. This is because most basic services such as PHC do not receive as much reimbursement neither do they charge user fees as other higher-level services.

*“, we can also overlook the services that don’t generate revenue. From an economic perspective, I’ll sit down as a facility and say if I invest in, for example, dialysis, SHA[Social Health Authority] will do 9,000 shillings per session, I’m supposed to do two sessions a week, in one year that’s 52 sessions. So, one patient for dialysis will give me 946,000 a year. Then I’m likely to say, we can invest in dialysis and leave immunization because immunization I’ll get 10 shillings. We are likely to also start to focus on services that give us more money, and we leave the ones that don’t give us more money.” National Government respondent*

Facilities have now started charging patient groups that were previously exempted from user fees. For example, while previously children under 5 years of age were exempt from user fees based on a ministry of health declaration, 26% of the facilities noted that under 5s were not exempt from user fees. This includes Kajiado, 17%, Kakamega, 44%, Kisumu, 19%, Lamu 33%, Migori, 15% and Nakuru, 27%. The basis of this is the lack of a policy document against charging user fees for under 5s. In addition, the respondents felt that in the long run once new finance bills are passed, then there will be an opportunity to amend prices, which may result in inequity. Fig 2 below shows providers’ perception on autonomy reforms increasing cost of service.

**Fig 2.**
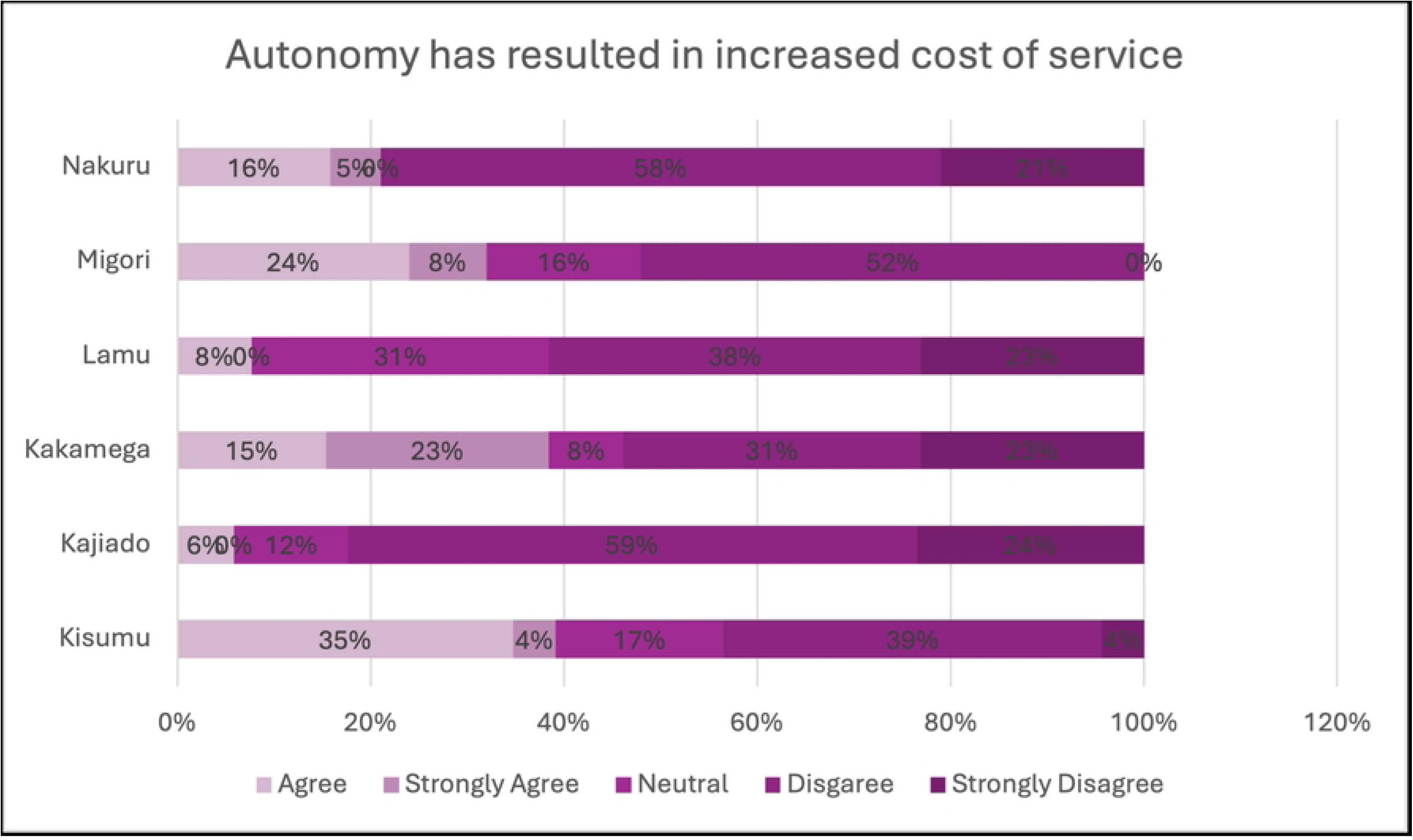
providers’ perception on whether autonomy has increased cost of services.

*“The good thing is that the finance bill regulates what facilities charge and the facilities cannot, don’t have the autonomy to set their own pricing charges away from what is in the finance bill and for the finance bill it has had public participation and all the actors coming in, so I wouldn’t say the facilities have that autonomy….They may probably influence in the long run where the amendments would be probably made in the finance bill, but until that is done then they don’t have that autonomy.” Sub County Manager County A*

Accountability over funds at the facility was a key issue raised by managers at both the national and county level. The county managers on the one hand felt that the health department were not keen on following the finance laws even when they were provided with the technical support

*“Then of course mismanagement. If we don’t take care of the money well, if the treasury teams now fail to build the capacity of the facilities to manage the money, then we can easily see some money getting lost.” National government respondent*

*“We have capacity. The only challenge is because I’m a finance person, I know, health is a very difficult department or the people there. Even if you were to take an accountant with capacity, they still feel they are more [laugh] important than these guys so, they don’t give them full support. They want to spend without accountability yet accountants are there to facilitate.” County Finance Manager county A*

Overall, the implementation of healthcare financing autonomy reforms as seen from the findings have positively enhanced managerial responsiveness and local accountability to some extent but remained and inconsistently implemented. Differences in legal interpretation and conceptualization, delays from the national treasury in funds disbursement and operational gaps have contributed to partial implementation public healthcare finance autonomy especially at primary health facilities. Autonomy is therefore partial, varying across functions and levels, with equity and sustainability concerns emerging.

## DISCUSSION

Acknowledging that facility autonomy reforms like all decentralization reforms are complex, and the impact may be mixed, depending on how they are implemented, (1,19), our study aimed to evaluate the emergence, implementation fidelity, and document the overall implementation experience of Kenya’s facility autonomy reforms.

Our analysis of the **reform’s emergence** revealed that the enactment of FIF laws at both national and subnational levels in Kenya was driven by political, technical, and public pressures. These demands arose primarily from bureaucratic obstacles in accessing funds, which undermined facility functionality, service readiness and, ultimately, service delivery. This led to public and technical teams’ complaints that called the politicians to action. Historical institutionalism played a crucial role in shaping both the reform’s trajectory and its implementation (20,21). While facility autonomy had previously existed in Kenya, it was significantly disrupted by devolution and the accompanying legal changes (3,4,13,22–25). However, technical experts in the counties—particularly early adopters—played a key role in sustaining the reforms, leveraging their prior experience with autonomy to advocate for its retention/reintroduction. These early adopters also facilitated peer learning, allowing other counties to observe and adopt best practices, thereby accelerating the reform’s expansion.

Politicians both at national and sub national levels played a key role in advocating for the reforms. A similar pattern was observed in Tanzania, where securing high-level political support was essential in advancing the reform (26). However, key differences emerged: while development partners played a leading role in shaping health financing reforms in pre-devolution Kenya and Tanzania, their role in this reform was more supportive than directive. This shift suggests a growing reliance on local expertise and governance structures in driving reforms forward (26,27).

Regarding **implementation fidelity**, our analysis revealed that all six counties deviated from certain provisions of both national and county-specific laws. Key areas of non-compliance included the retention of all funds at the source, allocation of matching funds from the exchequer, establishment of mechanisms to reimburse facilities for services provided to vulnerable populations, and granting full autonomy in financial management, procurement, and human resources. This limited decision space in some areas has also been shown in other countries including Fiji and Vietnam where health facilities were granted decision space, however decision making in human resources some financial aspects and procurement was limited (27–29).

Our findings also show that key implementation activities such as the development of regulations, training of managers, and incorporation of the facilities onto the IFMIS, were either partially done or not done at all. The limited training of managers on responsibilities that come with autonomy has been flagged as a major challenge influencing the implementation of autonomy reforms across several low-and middle-income countries including Indonesia, and Iran (27). Only routine preexistent structures such as supportive supervision and opening of bank accounts continued while incorporating autonomy. Also reinforcing historical institutionalism, that institutions sometimes resist change, and that change is gradual rather than radical (20,21). This partial implementation of policy activities may point to implementation capacity challenges and may compromise the full realization of the potential benefits of the reforms.

From literature, fidelity is determined by the complexity of the reform, facilitation support, the relevance of the reforms to the stakeholders, and relationship with moderators (30). While autonomy reforms were perceived as relevant across stakeholders, the intervention was complex, impacting several other laws, and entities. Some of the areas of noncompliance, particularly in retention of all funds at source, were influenced by operational needs at the county and sub-county levels, where delays in fund disbursement from the exchequer hindered their ability to function effectively. As a result, they diverted a portion of facility-generated funds to sustain their operations, despite being aware that these resources were already insufficient.

On addressing complexity of reforms, literature suggests that interventions that are explicitly explained are more likely to be implemented (30). While the laws described the what, regulations that ought to describe the how were not done, hence the significant gaps in implementation. Additionally, although subnational governments adapted the national and model laws to fit their local contexts, critical elements, such as reimbursement to facilities for providing healthcare to indigents, were overlooked in formulation of some county specific laws. This highlights a gap in policy formulation and the use of evidence, especially at the sub national level.

Our findings on the **implementation experience** of health facility autonomy reforms revealed a mix of enablers, challenges and unintended consequences. Key enablers included political support, support from development partners, institutional memory, technical capacity of the department of health, support from the county treasury and consensus among stakeholders. Barriers to autonomy reforms included resistance from county treasury, limited capacity for formulation of laws, and limited facility capacity to generate revenue. Unintended consequences included reduced exchequer allocations to the department of health and increased pressure for patients to pay all medical bills. Similarly, experiences from countries such as, Vietnam, Tanzania and Uganda show that autonomy reforms were politically charged, had both negative and positive outcomes and, in Vietnam’s case, the facility laws were changed multiple times indicating challenges in formulation of laws (26–28,31,32).

Finally, we found that even where autonomy reforms have been introduced, the extent of autonomy varied both within and between counties. The different levels of health facilities experienced autonomy differently, even when they were in the same county. The differences could be differences in how autonomy was operationalized given that none of the counties besides county E formed detailed regulations or guidelines to unpack the specific issues.

## CONCLUSION

This study has several limitations. First, the quantitative analysis relies on healthcare providers’ subjective perceptions of autonomy, meaning that some aspects may be shaped more by individual perspectives than by a standardized definition. Second, 5 of the 6 counties rolled out their laws prior to the national law, when evaluating fidelity, we use the national law and conceptual framework as gold standard and not each county specific law.

Despite its limitations, the study highlights three key priorities for strengthening health facility autonomy reforms. First, cunty governments should establish clear legal and policy frameworks that align autonomy with health system goals to ensure consistent implementation in line with health system goals. Second, to promote equity, the county government must reimburse primary healthcare services and healthcare care for vulnerable populations. Third, sustained investments are needed to build both the capacity of health facilities to deliver quality services and the leadership and management skills of facility managers and facility boards to fully realize the benefits of autonomy.

## Declarations

### Ethics approval and consent to participate

This study received ethics approval from the Scientific and Ethics Review Unit at Kenya Medical Research Institute, approval number KEMRI/SERU/CGMR-C/294/4708, and the National Commission of Science and Technology approval number NACOSTI/P/24/42187. We also sought approval from all institutions involved in the study.

## Consent for publication

Not applicable

## Availability of data and materials

The datasets generated and/or analysed during the current study are available in the Harvard Dataverse repository, Replication Data for: Examining the Implementation Process and Experience of health facility Autonomy Reforms in Kenya: A mixed methods study of counties in Kenya - Health Economics Research Unit (HERU) Dataverse

## Authors’ contributions

AMusiega: conceptualisation, methodology, formal analysis, funding acquisition, writing - original draft and project administration. JN: conceptualisation, methodology, formal analysis, writing - review and editing, supervision, funding acquisition and project administration. BA: conceptualisation, methodology, formal analysis, funding acquisition and writing - review and editing. HO: data collection, formal analysis and writing - review and editing. BM: conceptualisation, methodology, formal analysis and writing - review and editing. JN and BT: conceptualisation, methodology, formal analysis, writing - review and editing, supervision, funding acquisition and project administration. PMM: conceptualisation, methodology, supervision and funding acquisition. *EWangia^4^, KA, RR, JM SW, VT: data collection, interpretation of data, writing – review and editing.* EWong, CM, WN, AMusuva, FM and NR: conceptualisation, methodology, resources, writing - review and editing and supervision. EB: conceptualisation, methodology, formal analysis, resources, writing - review and editing, supervision and funding acquisition.

## Data Availability

The datasets generated and/or analysed during the current study are available in the Harvard Dataverse repository, https://dataverse.harvard.edu/dataset.xhtml?persistentId=doi:10.7910/DVN/SGINBJ

## Acknowledgements

We acknowledge the data collectors who collected the quantitative data - Jared Omondi, Hellen Amollo, Wycliffe Ochieng’ Tabela Akinyi, Erick Oloo, Annet Iminza, Babracks Chibole, Beth Makena Kiogora, Constant Mutanda, Cynthia Emali Mwakha, Edwin Nakitare, Fredrick Kagoni, Nicholas Sifuma, Yvonne Khakayi, Alice Chelsea Otieno, Amos Odhiambo Otieno, Norah Atieno Odhiambo, Loice Magembe Obaigwa, Florence Akinyi, Immaculate Nkasiti, Tina Sakimba, Leah Resiato, Catherine Mopia, Margaret Talian, Ann Silati, Esther Silantoi, Lucy Sintila, Nickson Sakimba, Enock Kuya, Farha Ahmed, Ernest Maina, Badru Hussein, Lucky Mzee, Vincent Chome, Nuru Salim, Rahma Said and Yvonne Ngalunja, Peninah Nasimiyu Machimbo, Peter Ondieki Nyamweya, Sharon Achieng Ondoro, Brian Kiboma Ondieki, Jemimah Bonareri Nyamwaro, Beth Wanjiku Kinyanjui, Hassan Ruto and Peninah Jebichii.

We also acknowledge the facility managers, from the 141 facilities and county managers from the six counties – Lamu, Nakuru, Kakamega, Kisumu, Kajiado, and Migori for accepting to be part of the study.

